# PREVALENCE, INCIDENCE AND DETERMINANTS OF QUANTIFERON-POSITIVITY IN SOUTH AFRICAN SCHOOLCHILDREN

**DOI:** 10.1101/2024.03.11.24304073

**Authors:** Justine Stewart, Neil Walker, Karen Jennings, Carmen Delport, James Nuttall, Anna K Coussens, Robin Dyers, David A Jolliffe, Jonathan C. Y. Tang, William D Fraser, Robert J Wilkinson, Linda-Gail Bekker, Adrian R Martineau, Keren Middelkoop

## Abstract

**Background:** Tuberculosis (TB) control requires the understanding and disruption of TB transmission. We describe prevalence, incidence and risk factors associated with childhood TB infection in Cape Town.

**Methods:** We report cross-sectional baseline and prospective incidence data from a large trial among primary school children living in high TB-burden communities. Prevalent infection was defined as QuantiFERON-TB Gold Plus (QFT-Plus) positivity as assessed at baseline. Subsequent conversion to QFT-Plus positivity was measured 3 years later among those QFT-Plus-negative at baseline. Multivariable logistic regression models examined factors associated with TB infection.

**Results:** QuantiFERON-positivity at baseline (prevalence: 22.6%, 95% Confidence Interval [CI]: 20.9 – 24.4), was independently associated with increasing age (adjusted odds ratio [aOR] 1.24 per additional year, 95% CI: 1.15 – 1.34) and household exposure to TB during the participant’s lifetime (aOR 1.87, 95% CI: 1.46 – 2.40). QFT-Plus conversion at year 3 (12.2%, 95% CI: 10.5-14.0; annual infection rate: 3.95%) was associated with household exposure to an index TB case (aOR 2.74, 95% CI: 1.05 to 7.18).

**Conclusion:** Rates of QFT-diagnosed TB infection remain high in this population. The strong association with household TB exposure reinforces the importance of contact tracing, preventative treatment and early treatment of infectious disease to reduce community transmission.

## BACKGROUND

Despite being treatable and curable, tuberculosis (TB) disease causes significant morbidity and mortality, impacting the lives of more than 10 million people every year, particularly in high-burden countries. Children under 15 years of age bear 11% of the global TB disease burden, with annual cases and deaths in this age group exceeding 1.1 million and 225,000 respectively^1^.

In high-burden countries *Mycobacterium tuberculosis* (*Mtb*) infection is often acquired in childhood^2^. TB infection prevalence, based on measures of *Mtb* antigen immune memory, has been reported to range from 21-41% among primary school children in high-burdened settings^3–7^. A substantial reduction in the incidence and mortality of TB requires the interruption of *Mtb* transmission^8^. Better understanding of the determinants of infection, would enable targeted medical, social and educational strategies to achieve TB control.

Factors positively associated with TB infection in high-burden settings include increasing age^4,9–12^, male sex^9–12^, known exposure to an infectious TB patient^10,11,13^, low vitamin D status^9,14^ and exposure to second-hand smoke^5,7,15^. There is inconsistency in risk factor findings between South African studies, such as the presence or absence of associations with socio-economic status^4,10^ and level of parental education^4,10^. Given the high prevalence of TB in South Africa, we aimed to determine the prevalence and incidence of QuantiFERON-TB Gold Plus (QFT-Plus) positivity as a measure of TB infection and identify associated risk factors among Cape Town primary school children.

## METHODS

This paper reports the analysis of cross-sectional baseline screening data and prospective incidence data from a large randomised placebo-controlled trial of Vitamin D supplementation for the prevention of TB infection among primary school children living in Cape Town, South Africa^16^. The study was performed in parts of Khayelitsha, Klipfontein and Mitchells Plain districts of Cape Town. The community has substantial unemployment, with high-density, typically poorly constructed dwellings and relies predominantly on public health and education services. The incidence of active TB in this area is estimated at 799/100 000^17^ population per annum, as compared to the South African rate of 468/100 000^18^.

Recruitment and screening have been described previously^19^. Potential participants were identified from 23 government-run primary schools within the districts described above. Eligibility criteria for the primary study included TB uninfected children (defined as a negative QFT-Plus interferon gamma release assay (IGRA) result at screening), in grade 1 to 4 and aged 6-11 years at enrolment. Written consent was obtained from the parent or legal guardian, followed by assent granted by the children. Exclusion criteria included previous active TB, positive IGRA or Mantoux test and/or treatment thereof. Children with known or suspected HIV infection were also excluded from the study.

At screening visits, data were collected by interviewer-led questionnaire, including socio-economic characteristics and medical details including history of exposure to a household TB case. Following assent, the children were examined for features suggestive of TB disease and the presence of a Bacillus Calmette–Guérin (BCG) scar. Eligible children underwent phlebotomy for QFT-Plus blood assay and serum 25-hydroxy-vitamin D (25[OH]D_3_) concentrations. The QFT-Plus samples were collected according to manufacturer’s specifications and were delivered to the laboratory in temperature-controlled and monitored portable incubators (Model NQ09, Darwin Chambers Company, MO, USA)^20^. All children who tested QFT-Plus positive or indeterminate were excluded from the prospective clinical trial after examination for active TB by a clinical team member. Eligible QFT-Plus negative participants were enrolled and received Vitamin D supplementation or placebo for up to three years, after which a repeat QFT-Plus assay was performed. Incident TB infection was defined as evidence of immune sensitisation based on QFT-Plus positive conversion at study completion.

### Laboratory methods

The latest generation IGRA was used to determine the outcome of interest. QFT-Plus has improved specificity and a lower indeterminate rate compared to the QuantiFERON-TB Gold In-Tube assay^21^. QFT-Plus specimens were processed at BARC-South Africa, a nationally accredited bioanalytical research laboratory. The assay was performed as per the manufacturers’ specifications in accordance with the FDA package insert. The interferon-gamma threshold of 0.35 IU/ml was used to determine a positive result. Serum concentrations of 25[OH]D_3_ were determined using liquid chromatography tandem mass spectrometry (LC-MS/MS)^19^ at a Bioanalytical Facility, University of East Anglia (Norwich, UK), as previously described^22^.

### Statistical Analysis

Statistical analysis was performed using Stata software (Version 17.0; StataCorp). Analysis was restricted to those with a valid QFT-Plus test result at the relevant time point. A conservative household crowding index was calculated based on the reported number of residents divided by the number of rooms in the house. Baseline (deseasonalised) and Year 3 25(OH)D concentrations were used. For both prevalent and incident data, univariate analyses explored factors predictive of QFT-positivity, employing Student’s t-tests and Wilcoxon sum rank tests for continuous variables and Chi-squared and Fisher’s exact tests for categorical variables, as appropriate. Annual infection rate was ARTI was estimated based on the QFT-conversion rate at follow-up using the formula: ARTI = 1 – [(1 - *P*)]^l/x^, where P = proportion of QFT-positive individuals at follow-up (12.2%), x = the mean follow-up time for individuals who returned a quantiferon result (3.22 yrs). Multivariable logistic regression models were developed to examine factors associated with prevalent and incident TB infection at baseline and Year 3 respectively. Multivariable analysis included all independent variables with a p-value of less than 0.1 in univariable analysis and were adjusted for age and study arm allocation. Separate models were developed to evaluate characteristics of household contacts associated with prevalent QFT-Plus status. These models were adjusted for age and school district, as the two primary predictive factors identified for QFT-Plus positivity in the main analysis.

The study received approvals from the University of Cape Town Faculty of Health Sciences Human Research Ethics Committee (Ref: 796/2015) and the London School of Hygiene and Tropical Medicine Observational/Interventions Research Ethics Committee (Ref: 7450-2).

## RESULTS

### Determinants of prevalent TB infection

Parents of 2852 children consented to their children’s study participation in the study (Figure 1). Overall, 40 (1.4%) children declined assent, or parents subsequently withdrew consent, and 211 (7.4%) children were ineligible. Reasons for ineligibility included difficult phlebotomy at baseline (84, 2.9%), family planning to move out of the study area within the study period (57, 2.0%) or history of previous latent or active TB (55, 2.0%).

**Figure 1:**
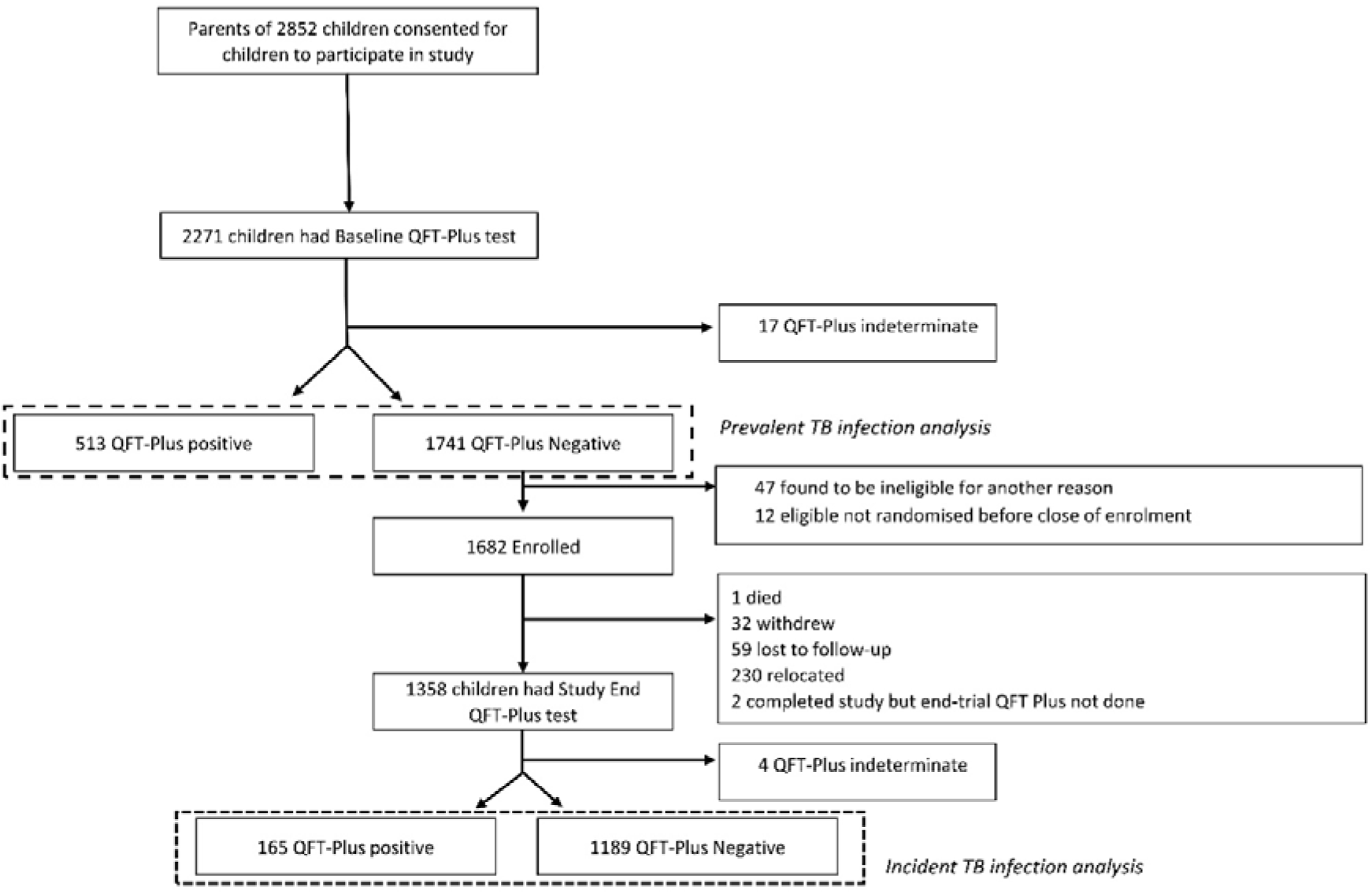
Participant flow. * Participants had ≥1 more reasons for ineligibility

In total, 2,271 children had a screening QFT-Plus assay, with 17 (0.7%) producing an indeterminate result. The cohort characteristics of the 2,254 participants with valid QFT-Plus results are outlined in Table 1. This cohort had a median age of 9.1 years (interquartile range [IQR]: 7.8-10.0) and 52.7% were female. Recruitment was spread throughout the study area with 800 (35.5%) participants from schools in Nyanga, 387 (17.2%) from Khayelitsha and 355 (15.8%) from Mitchells Plain areas. The median monthly household income of the cohort was 1,200 ZAR (IQR: 700-2500) with 52.4% living in informal housing. A total of 377 (16.6%) children had a history of exposure to 1 or more persons in their household living with pulmonary TB (PTB) during the child’s lifetime. Baseline serum 25(OH)D_3_ concentrations were available for 1,812 children; the mean concentration was 70.1 IU/ml (61.4 – 80.4) and 99 (5.5%) had a concentration of <50 nmol/L indicating vitamin D deficiency^23^.

**Table 1:**
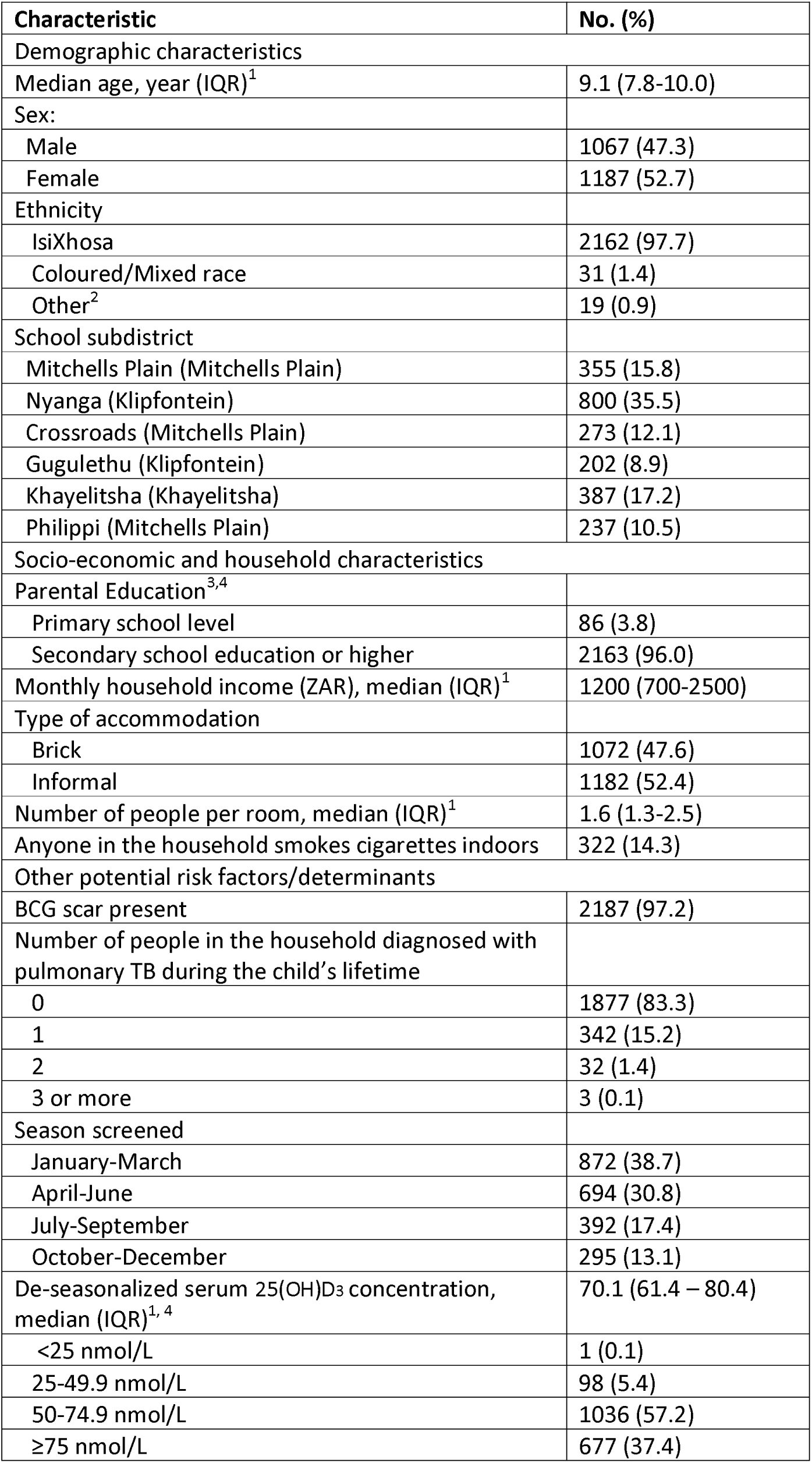
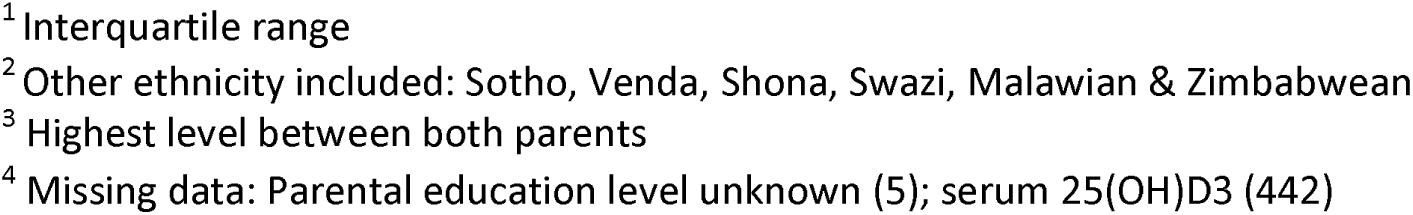
Characteristics of study participants included in analysis of determinants of baseline QFT-Plus status(n=2254)

| **Characteristic** | **No. (%)** |
| --- | --- |
| **Demographic characteristics** |  |
| Median age, year (IQR)<sup>1</sup> | 9.1 (7.8-10.0) |
| Sex: |  |
| Male | 1067 (47.3) |
| Female | 1187 (52.7) |
| Ethnicity |  |
| IsiXhosa | 2162 (97.7) |
| Coloured/Mixed race | 31 (1.4) |
| Other<sup>2</sup> | 19 (0.9) |
| School subdistrict |  |
| Mitchells Plain (Mitchells Plain) | 355 (15.8) |
| Nyanga (Klipfontein) | 800 (35.5) |
| Crossroads (Mitchells Plain) | 273 (12.1) |
| Gugulethu (Klipfontein) | 202 (8.9) |
| Khayelitsha (Khayelitsha) | 387 (17.2) |
| Philippi (Mitchells Plain) | 237 (10.5) |
| **Socio-economic and household characteristics** |  |
| Parental Education<sup>3,4</sup> |  |
| Primary school level | 86 (3.8) |
| Secondary school education or higher | 2163 (96.0) |
| Monthly household income (ZAR), median (IQR)<sup>1</sup> | 1200 (700-2500) |
| Type of accommodation |  |
| Brick | 1072 (47.6) |
| Informal | 1182 (52.4) |
| Number of people per room, median (IQR)<sup>1</sup> | 1.6 (1.3-2.5) |
| Anyone in the household smokes cigarettes indoors | 322 (14.3) |
| **Other potential risk factors/determinants** |  |
| BCG scar present | 2187 (97.2) |
| Number of people in the household diagnosed with pulmonary TB during the child's lifetime |  |
| 0 | 1877 (83.3) |
| 1 | 342 (15.2) |
| 2 | 32 (1.4) |
| 3 or more | 3 (0.1) |
| Season screened |  |
| January-March | 872 (38.7) |
| April-June | 694 (30.8) |
| July-September | 392 (17.4) |
| October-December | 295 (13.1) |
| De-seasonalized serum 25(OH)D<sub>3</sub> concentration, median (IQR)<sup>1, 4</sup> | 70.1 (61.4 – 80.4) |
| <25 nmol/L | 1 (0.1) |
| 25-49.9 nmol/L | 98 (5.4) |
| 50-74.9 nmol/L | 1036 (57.2) |
| ≥75 nmol/L | 677 (37.4) |

<sup>1</sup> Interquartile range
<sup>2</sup> Other ethnicity included: Sotho, Venda, Shona, Swazi, Malawian & Zimbabwean
<sup>3</sup> Highest level between both parents
<sup>4</sup> Missing data: Parental education level unknown (5); serum 25(OH)D3 (442)

The baseline prevalence of QFT-Plus-positivity in this cohort was 22.6% (95% Confidence Interval [CI]: 20.9 – 24.4). Factors associated with baseline QFT-Plus status are reported in Table 2. In univariate analysis, QFT-Plus positive status was positively associated with increasing age (odds ratio [OR] 1.21 per year added, 95% CI: 1.13-1.31), attending school in the Crossroads area (OR 1.91, 95% CI: 1.32 – 2.77), living in informal housing (OR 1.25, 95% CI: 1.03 – 1.52) and exposure to a household member living with PTB during the course of the participant’s lifetime (OR 1.93, 95% CI: 1.52 – 2.46). Further, school attendance in the Nyanga area (OR 1.31, 95% CI: 0.96 – 1.79) or the presence of anybody smoking cigarettes indoors trended towards a positive association with QFT-Plus positivity (OR 1.31, 95% CI: 1.00 - 1.71). In multivariable regression (Table 2), there remained a statistically significant, positive association of prevalent TB infection with increasing age (adjusted odds ratio [aOR] 1.24, 95% CI: 1.15 – 1.34), school attendance in the Crossroads area (aOR 1.54, 95% CI: 1.05 – 2.25) and household exposure to a person with PTB during the participant’s lifetime (aOR 1.87, 95% CI: 1.46 – 2.40).

**Table 2:**
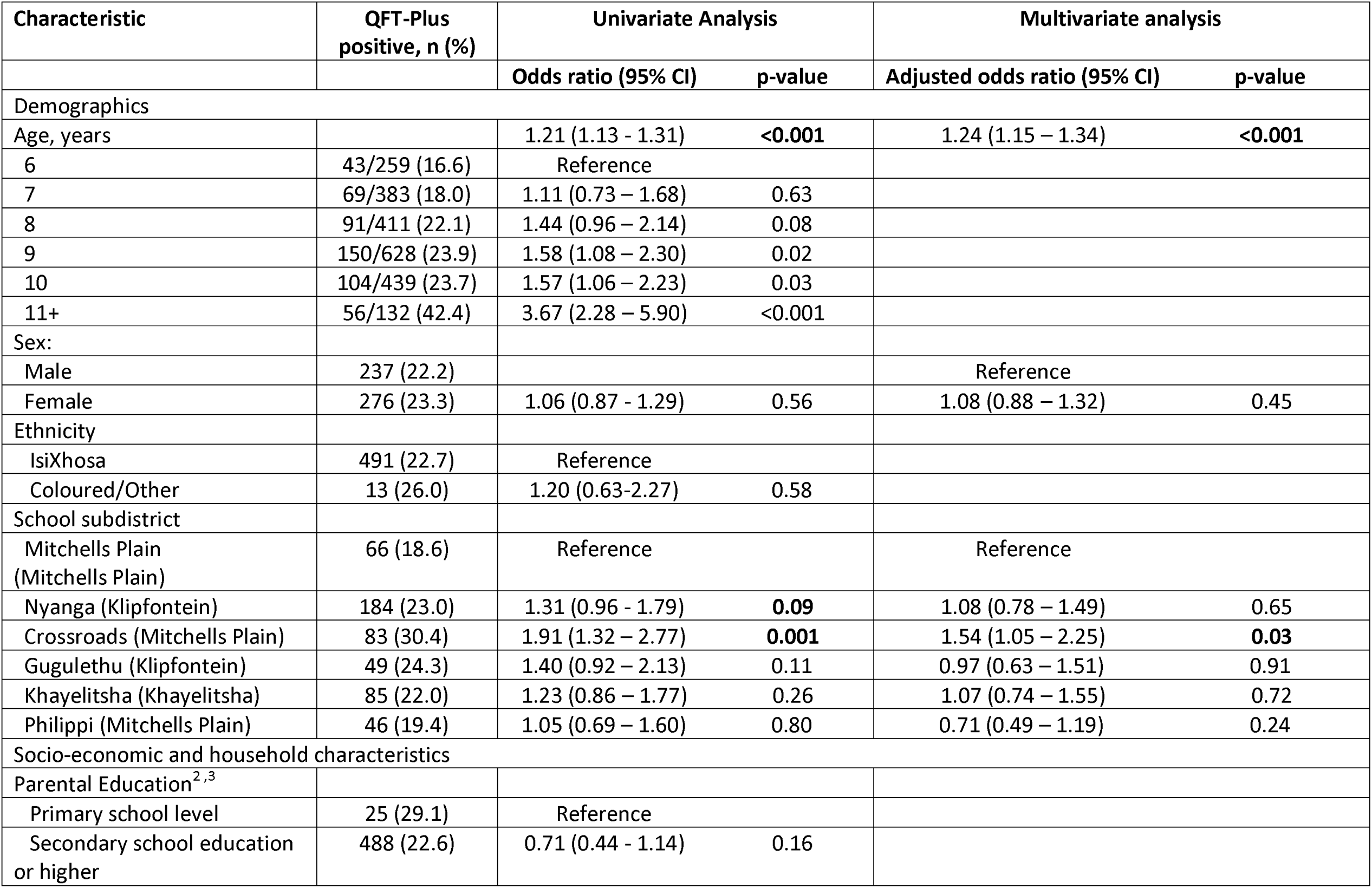
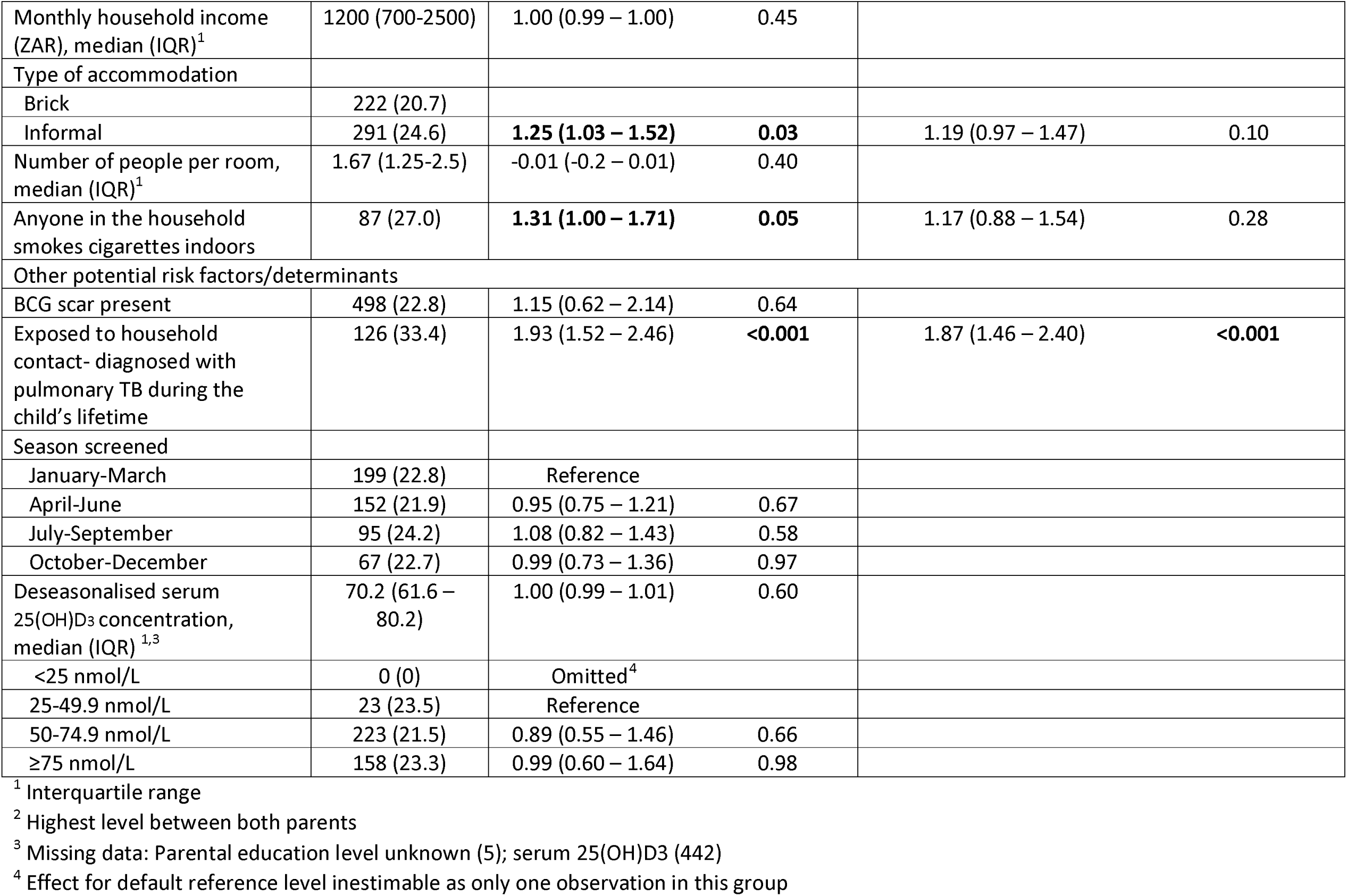
Determinants of prevalent QFT-Plus positivity (n=2254)

Among the 377 participants with a household PTB exposure in their lifetime, sleeping in the same room as the person with TB was the only statistically significant factor predictive of QFT-Plus positivity (Table 3). Neither time spent with the person with TB nor time from onset of cough to treatment initiation impacted participant QFT-Plus positivity.

**Table 3:** Determinants of infection risk in subset of participants reporting household exposure to PTB index case (n=377)

| Household TB contact: | QFT-Plus positive, n (%) | Univariate Analysis<sup>2</sup> |  |
| --- | --- | --- | --- |
|  |  | Odds ratio (95% CI) | p-value |
| Number of people in the household diagnosed with pulmonary TB during the child's lifetime |  |  |  |
| 1 | 115/342 (33.6) | Reference |  |
| 2 | 10/32 (31.3) | 0.93 (0.42 – 2.07) | 0.87 |
| 3 | 1/3 (33.3) | 0.96 (0.08 – 10.84) | 0.97 |
| Time since child's last contact with index case, median years (IQR)<sup>1</sup> | 1.26 (0-3.9) | 0.97 (0.89 – 1.05) | 0.39 |
| Child's proximity to index case |  |  |  |
| Sleeps in different room | 51/183 (27.9) | Reference |  |
| Sleeps in same room | 75/194 (38.7) | 1.79 (1.14 – 2.82) | **0.01** |
| Time child spent with index case |  |  |  |
| Not every day | 30/104 (28.9) | Reference |  |
| Every day, ≥50% of time | 51/131 (38.9) | 1.58 (0.90 – 2.77) | 0.11 |
| Every day, <50% of time | 45/142 (31.7) | 1.15 (0.66 – 2.03) | 0.62 |
| How long did index case cough before TB treatment was started (weeks) |  |  |  |
| ≤ 1 week | 12/35 (34.3) | Reference |  |
| 2 | 64/205 (31.2) | 1.0 (0.46 – 2.17) | 0.99 |
| 3 | 32/90 (35.6) | 1.16 (0.50 – 2.69) | 0.72 |
| 4 | 14/32 (43.8) | 1.64 (0.60 – 4.47) | 0.33 |
| ≥ 5 weeks | 4/15 (26.7) | 0.78 (0.20 – 3.04) | 0.72 |

<sup>1</sup> Interquartile range
<sup>2</sup> Adjusted for age and school subdistrict as factors, other than household contact, significantly associated with positive QFT-Plus

### Determinants of TB infection at study end

In the prospective analysis, among the 1,354 children who were QFT-Plus negative at baseline and who had a valid QFT-Plus results at the end of the study, 165 (12.2%; 95% CI: 10.5-14.0) had a positive end-study QFT-Plus result. This equated to an annual infection rate of 3.95% (95% CI – 3.36% to 4.54%). In both univariate and multivariate analysis of incident QFT-Plus positivity at the end of the study, exposure to a household TB contact over the 3 years of study participation was the only independently associated predictor (aOR 2.74, 95% CI: 1.05 to 7.18; Table 4). Female sex trended towards a significant association with incident QFT-Plus positivity in both univariate (p=0.08) and multivariate analysis (p=0.05). Factors associated with prevalent TB infection, age and area of school attendance, were not associated with incident infection in univariate analysis (p=0.13).

**Table 4:**
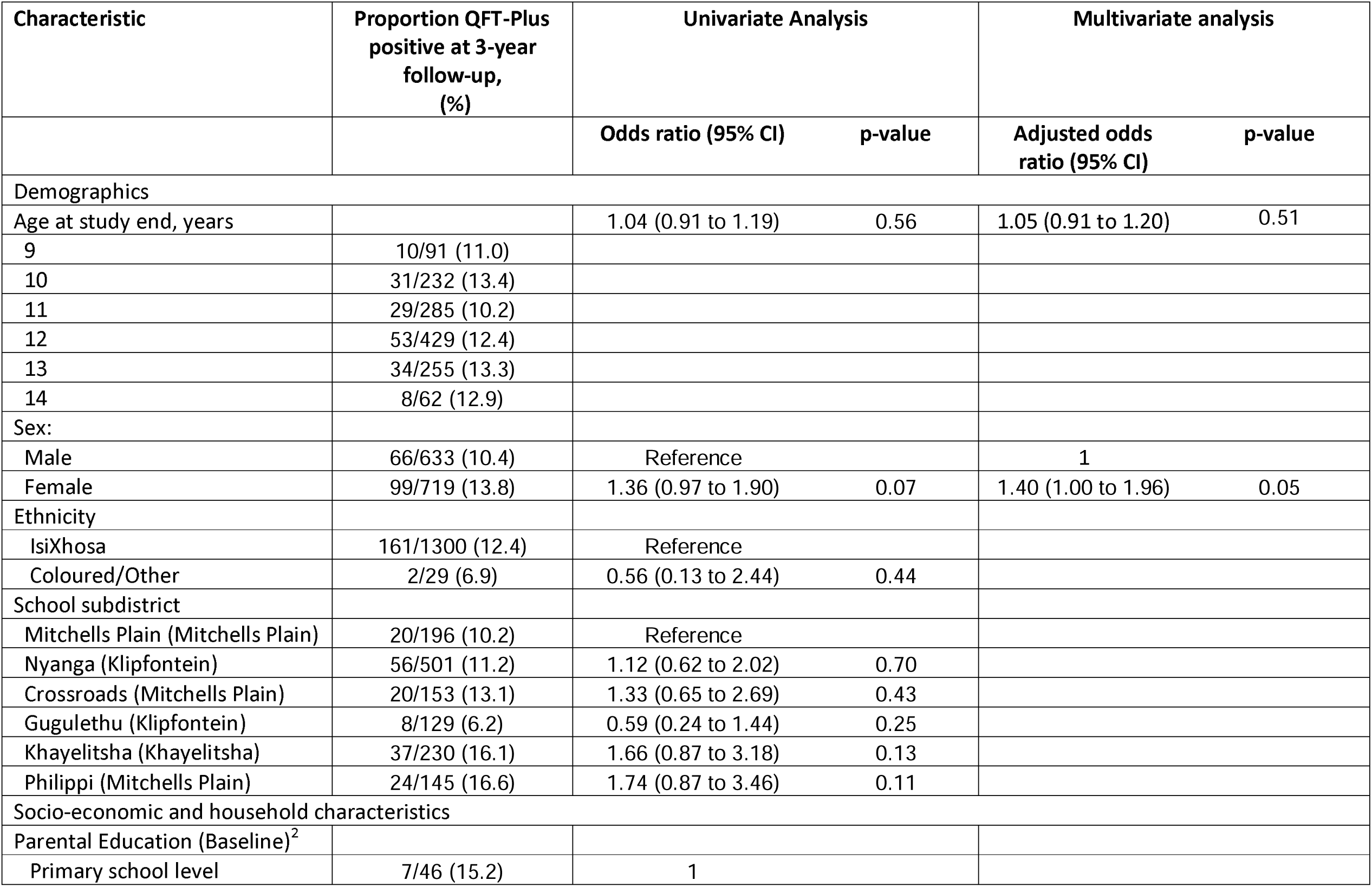
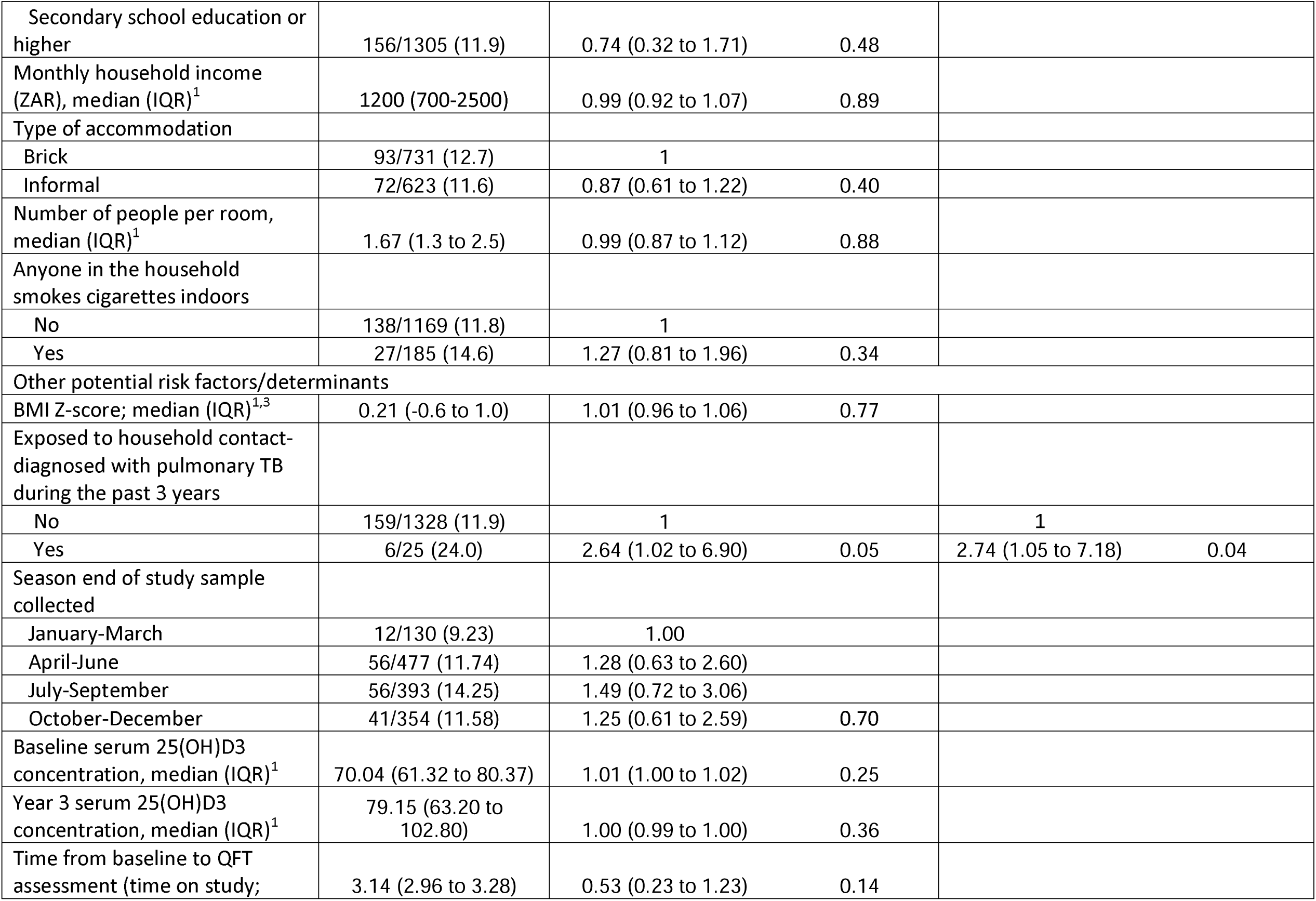
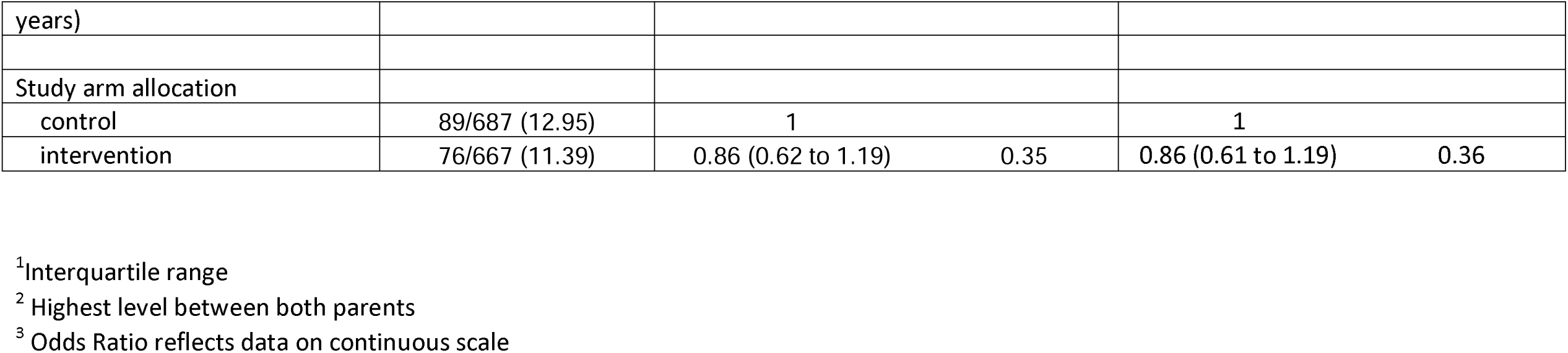
Determinants of incident QFT-Plus positivity (n=1354)

## DISCUSSION

This study reports a high prevalence and incidence of TB infection, defined as immune sensitisation QFT-Plus, among primary school children in a high-burden TB setting. The prevalence of 22.6% (95% CI: 20.9 – 24.4) is consistent with previously published data from similar demographic areas in Cape Town^3.10^ but is the first such data reported from the populous Klipfontein and Mitchells Plain districts and represents the most recent sub-Saharan TB infection prevalence data, utilising the improved diagnostic assay of QFT-Plus. Furthermore, 12% of children converted to QFT-Plus positivity, equating to a minimum annual infection rate of 3.95%, measured across the 3-year follow up period.

Key determinants of prevalent QFT-diagnosed TB infection in this study included increasing age, exposure to at least one person with PTB living in the same household over the lifetime of the child and the district in which the child attended school. Increasing age and household TB exposure are established risk factors for acquisition of TB infection^4,9–11,13^. Among participants who had lived with at least one person with PTB, only sleeping in the same room was statistically associated with prevalent TB infection, suggestive of a likely transmission link. Other anticipated factors, notably the number of persons who had TB in the house, and time spent with those with TB were not associated with baseline QFT-Plus positivity. This may be due to the lack of an infectious measure of index cases and the small number of participants included in this sub analysis, reducing power to detect associations.

Given limited scholar transport options in this district, the subdistrict of school attendance is a reasonable surrogate for the participants’ residential area. While particular subdistricts were more strongly associated with prevalent TB infection, there was no association between school subdistrict and incident infection. The association with prevalent TB infection may indicate historical differences between subdistricts such as variable timing of demographic and socio-economic expansion. However, the incident data suggests that the risk across areas is now homogenous.

In contrast to data recently published from a Mongolian cohort^9^, no association was identified between QFT-Plus positivity and vitamin D status or indoor cigarette smoking at home. This may be explained by the very discrepant prevalence of these factors between the populations: the South African cohort had an overall prevalence of vitamin D deficiency (25[OH]D <50 nmol/L) of 7.6%^19^ as compared to the Mongolian cohort prevalence of 95.6%^9^. Similarly, while smoking is a well-recognised risk factor for TB infection^15,24,25^, only 14.3% of parents reported that a household member smoked indoors, compared to 36.3% in the Mongolian study. The nature of exposure to cigarette smoke may also be different across the two settings: given the milder winters in Cape Town, smokers may be able to smoke outdoors more often when compared to smokers based in Mongolia with its more prolonged inclement winters.

The only measured factor associated with incident TB infection was exposure to a household TB case. Female participants trended towards a higher odds of incident infection compared to male participants. This is in keeping with other reports for this young adolescent age group^26^. This finding should be explored further in larger studies, together with possible explanations, such as different social mixing patterns between the sexes, or whether females spent more time in the household with a possible TB case^27,28^. The null association with pharmacological effect of Vitamin D supplementation in this clinical trial is an important finding which has been previously reported^16^, and is consistent with other recently reported work in young children^29^.

Key strengths of this study include the combination of cross-sectional and prospective data, enabling a rare opportunity to compare prevalent and incident TB infection risk factors in the same population, and the large sample of young school children. Schooling is compulsory in the age group studied and the implementation of this study across 23 government schools provided wide community representation, thus ensuring reasonable population generalisability. This study contributes to the few data evaluating IGRA testing as a marker for TB infection in a 6-14 year old paediatric population. The assay used was the latest generation of QFT tests, with improved diagnostic parameters and was performed by an accredited laboratory, with <1% of samples reported as “indeterminate”. Further, although the subset of participants reporting household exposure to a household person with PTB was relatively small, we obtained detailed information regarding household TB exposure, such as sleeping arrangements and the time spent together.

It is noted that this study sample is not a randomly selected cohort, but a screening cohort for a randomised placebo-controlled trial. This study cohort is largely limited to black South African children and is therefore not representative of all population groups. The association between prevalent TB infection and area of school attendance assumes that children attend school in their residential area, which is reasonable based on low economic resources and limited scholar transport options in the area. Furthermore, body mass index, a common factor associated with TB infection, could not be included in the prevalent infection analysis as these data were collected only on QFT-negative children eligible for the primary study.

In conclusion, TB infection remains a substantial burden in this setting. Our study adds original data from South Africa reporting a high prevalence and incidence of QFT-Plus diagnosed TB infection amongst the predominantly black South African primary school children enrolled in this study. The strong association with TB exposure in a household reinforces the importance of current WHO recommendations for contact tracing and early initiation of preventative or treatment regimens for all household contacts in high TB burden settings to avert or disrupt infectiousness of individuals, thus reducing household and community transmission of TB.

## Data Availability

Anonymised data may be requested from the corresponding author to be shared subject to terms of research ethics committee approval.

## ACKNOWLEDGEMENTS

We thank all the children who participated in the study, and their parents / guardians. We also thank the Western Cape Education Department and the schools that supported this work. This research was funded by the UK Medical Research Council (refs MR/R023050/1 and MR/M026639/1, both awarded to ARM). RJW was supported by Wellcome (104803, 203135). He also receives support from the Francis Crick Institute which is funded by Cancer Research UK (FC2112), the UK Medical Research Council (FC2112) and Wellcome (FC2112).

